# ASSOCIATION BETWEEN NUTRITIONAL STATE AND PHASE ANGLE IN SYMPTOMATIC AND ASYMPTOMATIC HTLV-1 INFECTED PATIENTS

**DOI:** 10.1101/2022.03.22.22272740

**Authors:** Adriana Bacelo, Abelardo Araújo, Paloma Torres, Patrícia Brito, Cristiane Almeida, Clévio Fonseca, Marcel Quintana, Cláudia dos S. Cople-Rodrigues, Pedro E. A. A. do Brasil, Naíse Rocha

## Abstract

**Introduction:** The nutritional status of symptomatic and asymptomatic human T-cell lymphotropic virus type 1 (HTLV-1) infected patients is understudied. The phase angle (PA) has been described in the scientific literature as a prognostic indicator of nutritional status, but this has not been sufficiently discussed in the literature. Therefore, neither the impact of the infection nor the disease’s progression is sufficiently known regarding the nutritional status, body condition or composition.

**Objective:** To compare the nutritional status of symptomatic and asymptomatic adult individuals infected by HTLV-1, using the PA and anthropometric measures as a prognostic indicator in the HTLV-1 infected population.

**Methodology:** This was an observational, cross-sectional study with symptomatic and asymptomatic HTLV-1 patients followed up at the Neurology outpatient clinic of the Evandro Chagas National Institute of Infectious Diseases (INI-FIOCRUZ), Brazil, from September 2015 to September 2019. Anthropometric measures and indices (body weight, height, body mass index-BMI, mid-upper arm circumference-MUAC, triceps skinfold - TSF, and mid- arm muscle circumference-MUAC), and bioimpedance (phase angle-PA, percentage of lean mass-%LM, and percentage of fat mass-%FM) were checked to assess the nutritional status. Anthropometric variables were classified according to reference values and compared between groups (symptomatic and asymptomatic). Individuals were considered malnourished when at least one of the nutritional assessment results was outside the reference values. PA was correlated with the nutritional status, and groups of symptomatic and asymptomatic were compared to each other. The R-project® program version 3.0.2 was used to analyze the data. Differences were considered significant when the p-value was ≤ 0.05.

**Results:** Ninety-one patients were evaluated, 33 (36.3%) asymptomatic and 58 (63.7%) symptomatic. The majority were female (61.5%) and the median age was 60 (55-58) years. Symptomatic participants, compared to asymptomatic, had a lower proportion of overweight/obesity (51.7% vs 78.8%; p =0.0171), lower BMI (25.47 ± 5.06 kg/m^2^ vs 30.08 ± 5.61 kg/m^2^; p = <0.001), MUAC (29.56 ± 5.13 cm vs 33.22 ± 4.21 cm; p =0.0011), and %FM (30.75% vs 36.60%; p =0.0064), however, had a higher %LM (68.95% vs 63.40%; p =0.0299). All participants presented PA, however there was no difference between symptomatic (5.74º ± 1.18) and asymptomatic (6.21º ± 1.16).

**Conclusion:** Overweight and obesity were prevalent, especially among asymptomatic participants. Symptomatic participants had lower BMI, MUAC and %FM. Mid-upper arm circumference was considered a good parameter for monitoring the nutritional status of people with HTLV, mainly in situations where weight measurement is not viable. PA was altered in both groups, therefore, it cannot be used as a disease progression indicator, but it does indicate that HTLV infection alone should be a risk of cellular membrane integrity damage. Studies using PA assessment in HTLV-1 carriers are needed.

**AUTHOR’S SUMMARY:** HTLV is a disease very little explored, and in the scientific field of nutrition it is no different, we found few studies that associate this population and their nutritional status. In academic literature we can find the association of weight, height and BMI, rare studies with bioimpedance assessments.

Until this moment, no study has associated nutritional status with the phase angle, which is being used in several infectious diseases as a prognostic indicator of cell membrane integrity.

In this study, we note that even though the phase angle values are not significant, they show that, regardless of the symptoms, patients who are infected with the HTLV virus are already considered to have damage to the membrane integrity, which makes us emphasize the importance of new studies to a better understanding of factors related to weight gain and probable nutritional deficiencies.

## INTRODUCTION

In 1980 the T-cell lymphotropic virus (HTLV) was the first isolated human retrovirus and was subsequently classified: as HTLV-1 and HTLV-2 (1). HTLV is a neglected condition in such a way that it is not in the World Health Organization (WHO) list of neglected diseases and it is not considered in the surveillance (2–4). It’s very low frequency of clinical manifestations is one of the main reasons why HTLV infection is a neglected condition. Recent data suggests that only 5% of the HTLV-1 infected patients are symptomatic (3,5).

Nevertheless, current estimation of HTLV-1’s burden is approximately 20 million of infected people worldwide (6). HTLV can be divided into two types: HTLV-1 and HTLV-2, and HTLV-1 is the most prevalent. HTLV-1 is related to a higher occurrence of adult T-cell leukemia/lymphoma (ATLL) and it is also associated with myelopathy/tropical spastic paraparesis (HAM/TSP). HAM/TSP is a chronic (lifelong) and slow progressive neurological condition (7,6,3).

There is a health-disease binomial directly related to conditions that have an impact on nutritional status, which in turn will impact immune status (8,9). Additionally, nutritional issues, such as overweight/obesity, underweight, malnutrition and micronutrient deficiency are a significant public health problem in developing countries or in poor populations around the world. These nutrition issues may have important repercussions on quality of life (10,11).

Nutritional status is relevant in every clinically ill patient. Alongside anthropometric biomarkers, bioelectrical impedance analysis (BIA) is useful to assess nutritional status in clinical practice. The phase angle (PA), one of BIA’s parameters, is related to cell function and integrity and there is evidence pointing to its ability to be a prognostic marker in some health conditions: such as in critically ill patients, in Crohn’s disease, oncologic, cirrhotic, HIV patients and various other conditions (12–15).

Lower PA values represent low reactance (Xc) and high resistance (R), which can be associated with the presence of a health condition or more severe clinical manifestations, cell death, or some change in the selective membrane permeability. On the other hand, higher values represent high Xc and low R and may be associated with a more significant amount of intact cell membranes, that is, greater body cell mass, and with a good state of health (16). However, the phase angle’s reference values and decision thresholds are still debatable and there is no established universal normal range (17).

The present study aimed to compare symptomatic and asymptomatic adults infected with HTLV-1 regarding their PA and nutritional status in order to discuss the potential use of PA as a prognostic marker.

## METHODS

### Study design

This was a cross-sectional study conducted at the outclinic of Evandro Chagas National Institute of Infectious Disease (INI-Fiocruz), Rio de Janeiro city, Brazil. The patients referenced to INI-Fiocruz or seeking medical attention regarding HTLV infection at INI-Fiocruz were submitted to a diagnostic investigation including an ELISA test with the anti HTLV I / II SYM Solution® kit. If there was confirmation of HTLV infection from the serological test, an additional polymerase chain reaction (nested PCR) test was performed. Standardized medical attention and follow-up by the neurologists was offered to these patients at the outclinic.

From the patients already in follow-up at the institute, the study screened and recruited from September 2015 to September 2019. All patients included met the following eligibility criteria: age of 19 or older; serological evidence of HTLV, with additional positive nested PCR; signed written Informed Consent Form (ICF). The exclusion criteria were: concurrent conditions other than HTLV infection that could impact the nutritional status such as hypermetabolic, malabsorptive, or immunosuppressive clinical conditions (HIV, cancer, intestinal parasites, Crohn’s disease, pregnancy); use of chemotherapy in the previous six months; protein, caloric or protein-calorie nutritional supplementation in the previous six months.

### Nutritional status and phase angle

The following anthropometric parameters were performed in the nutritional assessment: body weight, height, arm circumference (AC), triceps skinfold (TSF) and mid-arm muscle circumference (MUAC). Gender and age were obtained to classify anthropometric parameters according to the cutoff points established at the Food and Nutrition Surveillance System (SISVAN) (18). Body weight and height measurements were used to determine the Body Mass Index (BMI) as proposed by Lohman (16) and adopted by the Ministry of Health (19) and the World Health Organization (20).

BIA was performed by the “Biodynamics BIA 450”. It determines the body composition by an alternating electrical current of low intensity (500 to 800 MA) and high frequency (50KHZ), which travels through the body tissues and establishes the body mass distribution through the measurement of tissue counter-positives. The PA was measured in BIA by calculating the resistance: range from 200 to 1500 ohms, resolution 0.1 ohm and accuracy 0.1 percent; reactance: Range from 0 to 300 ohms, resolution 0.1 oh and accuracy 0.2 percent; phase Angle: range 0 to 20 degrees, resolution 0.1-degree, accuracy 0.2 percent (Xc/R) x180 °/π) by calculating Impedance (Z), the sum of the resistance (R) and Xc arc tangent (Xc/R) x180 °/π).

Before the BIA test, the team instructed participant to: discontinue the use of diuretic drugs for 24 hours, be fasting for 4 to 6 hours, be in alcoholic withdrawal for at least 24 hours, avoiding the consumption of foods rich in caffeine (coffee, dark teas, cola-based soft drinks, chocolate) for at least 24 hours, be out of menstrual period, and have not had a fever in the previous 24 hours, nor have practiced intense physical activity in the same period. The team also asked the participant to urinate minutes prior to the test. The researcher also emphasized that the patients were instructed to attend the neurologist’s appointment in fasting for blood tests. BIA was not performed if the patient had a pacemaker, metal plates around the body, or was pregnant.

To perform the BIA, the participant was asked to previously remove all metallic adornments before lying on a stretcher in a horizontal position at zero inclination (supine position). Then four electrodes (tetrapolar model) were positioned on the same side of the body - two in the hand close to the metacarpal-phalangeal articulation of the dorsal surface on the wrist, between the distal prominences of the radius and the ulna, and two on foot, in the transverse arch of the upper surface and on the ankle, between the lateral and medial malleoli. All participant information necessary for the examination, such as age, sex, weight, height, the habit of physical activity, including its intensity and frequency, were inserted in the equipment.

The BIA has been identified as a more accurate method for assessing body composition because, in addition to the description of body compartments, it informs the PA (14).

PA is considered a prognostic indicator for critically ill patients because it is related to membrane integrity (21,22).

### Analysis plan

For descriptive analysis, categorical variables will be presented as counts and frequencies, and continuous variables will be presented as mean variables (and standard deviation) or median (and interquartile intervals) as appropriate. Dispersion measures of PA and other nutritional biomarkers (LM%, FM%, weight, height, BMI, AC, TSF and MUAC) were estimated among the HTLV clinical status, stratified by age and sex. Whenever variables are compared between symptomatic and asymptomatic HTLV groups, the hypothesis tests indicated in the results are used. Fisher, t-student test or Wilcoxon test for continuous variables were used. For the variables already categorized, the chi-square test was evaluated. At the end, an ordinary least square regression was condudcted to adjust the effect of HTLV symptoms for age and sex on PA. Data were analyzed using the program R-project®.

### Ethical aspects

The present work was approved by the National Institute of Infectious Diseases (INI-FIOCRUZ) Research Ethics Committee (CAAE 16714219.4.0000.5262 on 10/25/2019).

## RESULTS

One hundred and ten patients were recruited, 19 were excluded (17%), as they did not meet the inclusion criteria, leaving a total of ninety-one participants with HTLV-1 were included, 58 (63.7%) were symptomatic and 33 (36.3%) were asymptomatic. Participants had a median age of 60 years, and were predominantly female. About 62% had BMI values indicating obesity or overweight, and BMI was higher among the asymptomatic group. (Table 1). Symptomatic group also had lower values of MUAC, and %FM, however, and higher values of %LM (Table 2).

**Table 1.**
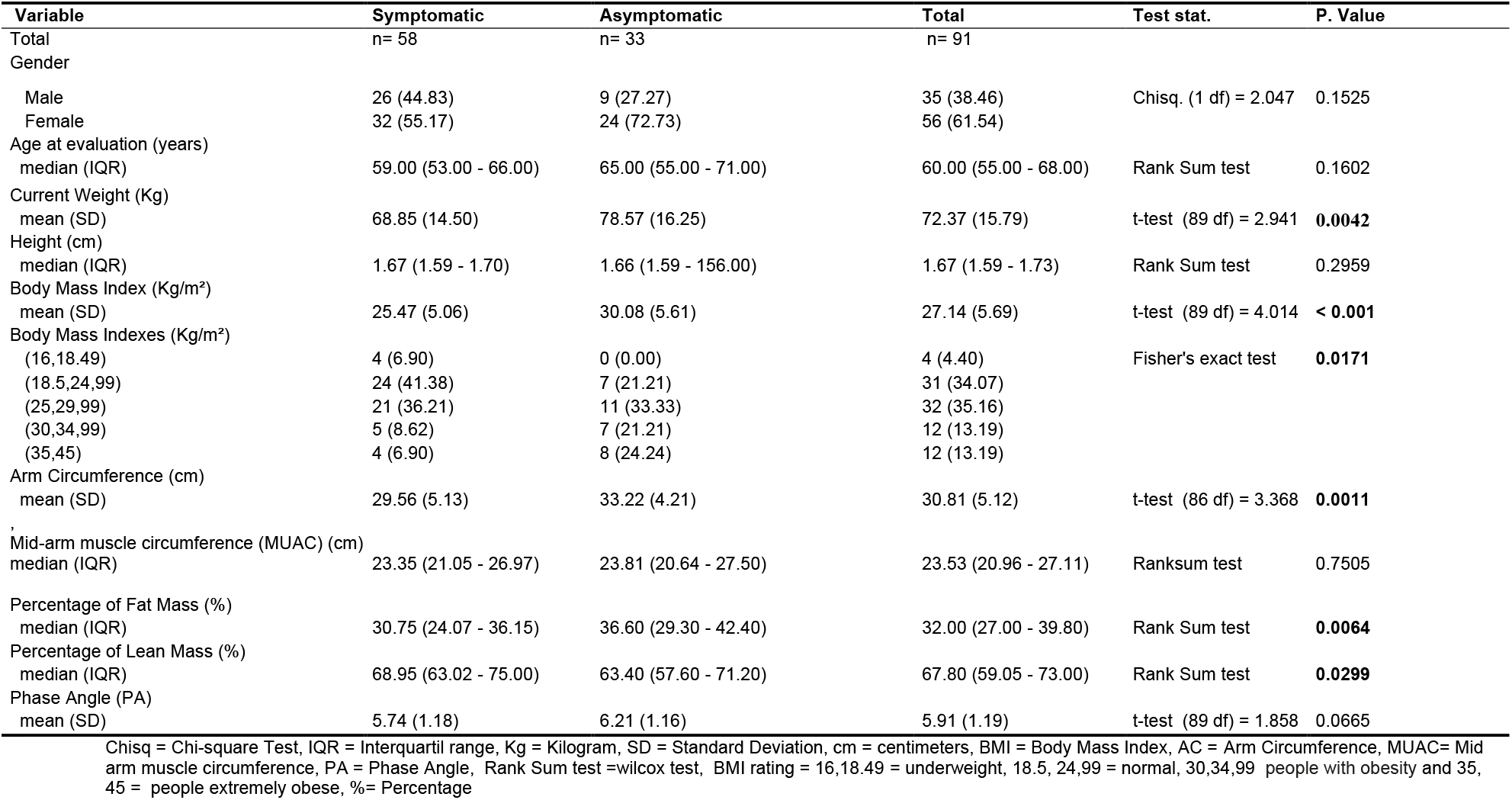
Descriptive characteristics of the HTLV population

| Variable | Symptomatic<br>n= 58 | Asymptomatic<br>n= 33 | Total<br>n= 91 | Test stat. | P. Value |
| --- | --- | --- | --- | --- | --- |
| Total |  |  |  |  |  |
| Gender |  |  |  |  |  |
| Male | 26 (44.83) | 9 (27.27) | 35 (38.46) | Chisq. (1 df) = 2.047 | 0.1525 |
| Female | 32 (55.17) | 24 (72.73) | 56 (61.54) |  |  |
| Age at evaluation (years) |  |  |  |  |  |
| median (IQR) | 59.00 (53.00 - 66.00) | 65.00 (55.00 - 71.00) | 60.00 (55.00 - 68.00) | Rank Sum test | 0.1602 |
| Current Weight (Kg) |  |  |  |  |  |
| mean (SD) | 68.85 (14.50) | 78.57 (16.25) | 72.37 (15.79) | t-test (89 df) = 2.941 | **0.0042** |
| Height (cm) |  |  |  |  |  |
| median (IQR) | 1.67 (1.59 - 1.70) | 1.66 (1.59 - 1.56.00) | 1.67 (1.59 - 1.73) | Rank Sum test | 0.2959 |
| Body Mass Index (Kg/m<sup>2</sup>) |  |  |  |  |  |
| mean (SD) | 25.47 (5.06) | 30.08 (5.61) | 27.14 (5.69) | t-test (89 df) = 4.014 | **< 0.001** |
| Body Mass Indexes (Kg/m<sup>2</sup>) |  |  |  |  |  |
| (16,18.49) | 4 (6.90) | 0 (0.00) | 4 (4.40) | Fisher's exact test | **0.0171** |
| (18.5,24,99) | 24 (41.38) | 7 (21.21) | 31 (34.07) |  |  |
| (25,29,99) | 21 (36.21) | 11 (33.33) | 32 (35.16) |  |  |
| (30,34,99) | 5 (8.62) | 7 (21.21) | 12 (13.19) |  |  |
| (35,45) | 4 (6.90) | 8 (24.24) | 12 (13.19) |  |  |
| Arm Circumference (cm) |  |  |  |  |  |
| mean (SD) | 29.56 (5.13) | 33.22 (4.21) | 30.81 (5.12) | t-test (86 df) = 3.368 | **0.0011** |
| Mid-arm muscle circumference (MUAC) (cm) |  |  |  |  |  |
| median (IQR) | 23.35 (21.05 - 26.97) | 23.81 (20.64 - 27.50) | 23.53 (20.96 - 27.11) | Ranksum test | 0.7505 |
| Percentage of Fat Mass (%) |  |  |  |  |  |
| median (IQR) | 30.75 (24.07 - 36.15) | 36.60 (29.30 - 42.40) | 32.00 (27.00 - 39.80) | Rank Sum test | **0.0064** |
| Percentage of Lean Mass (%) |  |  |  |  |  |
| median (IQR) | 68.95 (63.02 - 75.00) | 63.40 (57.60 - 71.20) | 67.80 (59.05 - 73.00) | Rank Sum test | **0.0299** |
| Phase Angle (PA) |  |  |  |  |  |
| mean (SD) | 5.74 (1.18) | 6.21 (1.16) | 5.91 (1.19) | t-test (89 df) = 1.858 | 0.0665 |

Chisq = Chi-square Test, IQR = Interquartil range, Kg = Kilogram, SD = Standard Deviation, cm = centimeters, BMI = Body Mass Index, AC = Arm Circumference, MUAC= Mid arm muscle circumference, PA = Phase Angle, Rank Sum test =wilcox test, BMI rating = 16,18.49 = underweight, 18.5, 24,99 = normal, 30,34,99 people with obesity and 35, 45 = people extremely obese, %= Percentage

**Table 2.** Anthropometry biomarkers according to HTLV population symptoms, sex and age.

| Variable | Sex<br>Female /<br>Male | Age<br>Years | Symptomatic<br>Mean ±SD (min,max) [N] | Asymptomatic<br>Mean ±SD (min,max) [N] | Total<br>Mean ±SD (min,max) [N] | p valor |
| --- | --- | --- | --- | --- | --- | --- |
| **Arm circumference (AC - cm)** | Female | 27 - 59 | 28.62 ±5.37 (24.70,31.38) [16] | 37.90 ±2.23 (36.75,39.50) [8] | 31.44 ±6.33 (27.75,36.75) [24] | **0.000736** |
| Female - normal (23.0 - 38.0cm) | Female | 60 - 81 | 27.82 ±3.81 (24.38,29.85) [16] | 31.74 ±3.36 (29.25,34.75) [15] | 29.42 ±4.07 (27.00,32.15) [31] | **0.015465** |
| Male - normal (26.2 - 36.9cm) | Male | 27 - 59 | 31.18 ±6.82 (26.00,35.90) [14] | 32.12 ±2.69 (30.88,33.25) [4] | 31.40 ±6.03 (27.00,35.50) [18] | 0.820739 |
| | Male | 60 - 81 | 31.63 ±3.83 (29.80,33.95) [12] | 32.45 ±5.30 (29.48,34.62) [5] | 31.85 ±4.08 (29.80,34.05) [17] | 0.851282 |
| **Mid-arm muscle circumference (MUAC- cm)** | Female | 27 - 59 | 23.00 ±3.51 (20.99,25.48) [16] | 22.28 ±9.25 (20.99,28.10) [8] | 22.76 ±5.85 (20.99,25.80) [24] | 0.528 |
| Female - normal (18.8 - 26.6cm) | Female | 60 - 81 | 22.00 ±2.75 (20.81,23.27) [16] | 17.88 ±10.78 (10.82,24.34) [15] | 20.01 ±7.90 (20.49,23.89) [31] | 0.828 |
| Male - normal (23.5 - 31.8cm) | Male | 27 - 59 | 23.71 ±8.70 (19.93,29.31) [14] | 25.70 ±1.47 (24.90,25.83) [4] | 24.15 ±7.68 (21.28,28.16) [18] | 0.645 |
| | Male | 60 - 81 | 25.09 ±8.15 (25.42,27.76) [12] | 21.50 ±11.91 (21.35,27.50) [5] | 24.04 ±9.16 (25.01,27.50) [17] | 1.000 |
| **Tricipital skinfold (TSF - mm)** | Female | 27 - 59 | 17.91 ±7.80 (12.07,22.67) [16] | 35.17 ±17.14 (31.45,41.05) [8] | 23.66 ±14.08 (13.78,32.40) [24] | **0.00451** |
| Female - normal (12.0 - 39.0mm) | Female | 60 - 81 | 18.53 ±8.09 (13.43,23.95) [16] | 18.30 ±11.92 (9.30,26.60) [15] | 18.42 ±9.95 (13.16,25.75) [31] | 0.59318 |
| Male - normal (05.0 - 20.8mm) | Male | 27 - 59 | 17.01 ±6.91 (13.35,22.07) [14] | 20.10 ±9.98 (12.62,25.42) [4] | 17.69 ±7.47 (12.70,22.85) [18] | 0.79052 |
| | Male | 60 - 81 | 12.75 ±7.22 (8.33,18.20) [12] | 15.04 ±10.81 (8.90,18.00) [5] | 13.42 ±8.14 (8.40,18.10) [17] | 0.83293 |
| **Body mass index (BMI - Kg/m<sup>2</sup>)** | Female | 27 - 59 | 26.43 ±6.69 (21.88,30.80) [16] | 36.30 ±3.24 (34.50,38.42) [8] | 29.72 ±7.41 (22.66,36.03) [24] | **0.00297** |
| Adult - normal (18.5 - 24.9Kg/m<sup>2</sup>) | Female | 60 - 81 | 24.07 ±4.51 (20.42,26.80) [16] | 27.21 ±4.62 (23.35,30.15) [15] | 25.59 ±4.76 (21.95,28.80) [31] | 0.07525 |
| Elderly - normal (23.0 - 28.0Kg/m<sup>2</sup>) | Male | 27 - 59 | 26.20 ±4.99 (23.46,28.04) [14] | 29.35 ±4.50 (26.93,30.92) [4] | 26.90 ±4.94 (24.17,29.12) [18] | 0.23268 |
| | Male | 60 - 81 | 25.21 ±3.10 (24.75,26.89) [12] | 30.02 ±5.62 (25.60,31.70) [5] | 26.62 ±4.43 (25.21,28.30) [17] | 0.08177 |

AC= Arm circumference, MUAC= Mid-arm muscle circumference BMI = Body mass index, Max= Maximum, Min=Minimum, N= sample size, SD = Standard Deviation, TSF =Tricipital skinfold; p value for the wilcox test ; Source: AC, AMC and TSF - Frisancho (1990) (39) , BMI Adult - Who (1995) (40), BMI Seniors - OPAS (2002) (41)

Regarding the anthropometric measures, differences can be observed among the symptomatic and asymptomatic group in some specific strata, such as differences between the AC within the female participants, and differences between the TSF and BMI within the younger females’ participants (Table 2). And even in the AC regardless of gender (figure 1).

**Figure 1.**
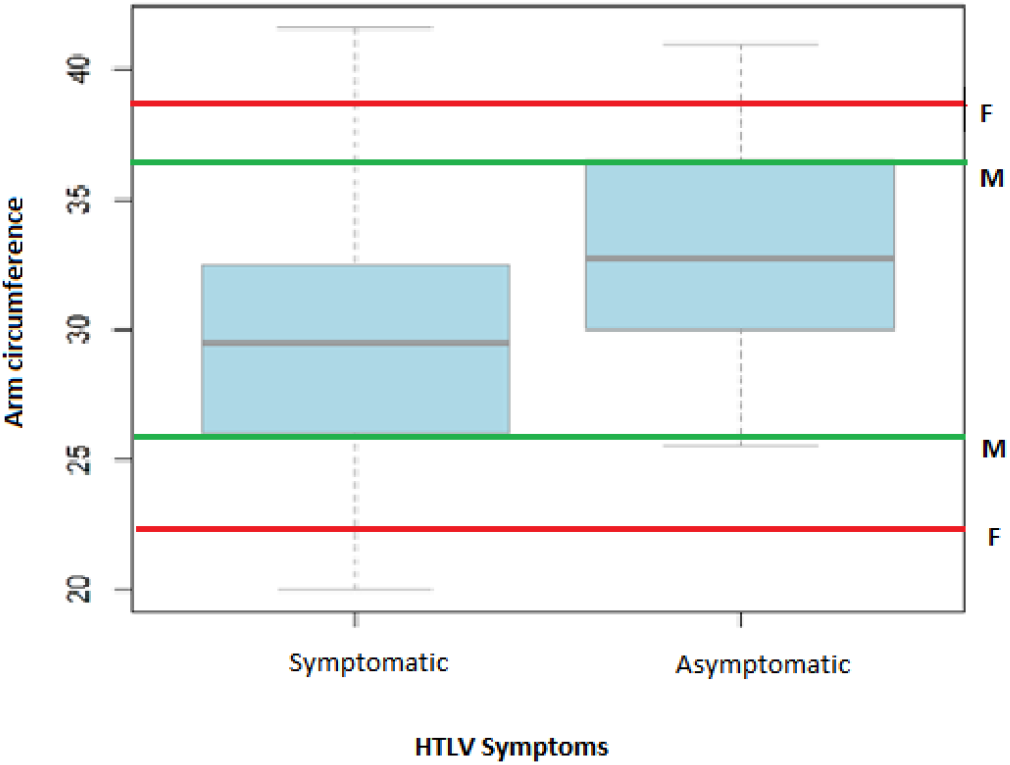
Box plot of arm circumference comparison of symptomatic and asymptomatic HTLV participants. HTLV = Human T-cell Leukemia-lymphoma Virus, F = female normal range amplitude limits, M = male normal range amplitude limits

The majority of symptomatic and asymptomatic had MUAC values within the normal range, regardless of gender. AC seems to be positively correlated with MUAC and BMI. (Figure 2). Additionally, asymptomatic tend to have higher AC values when MUAC is kept fixed in lower values when compared to symptomatic.

**Figure 2.**
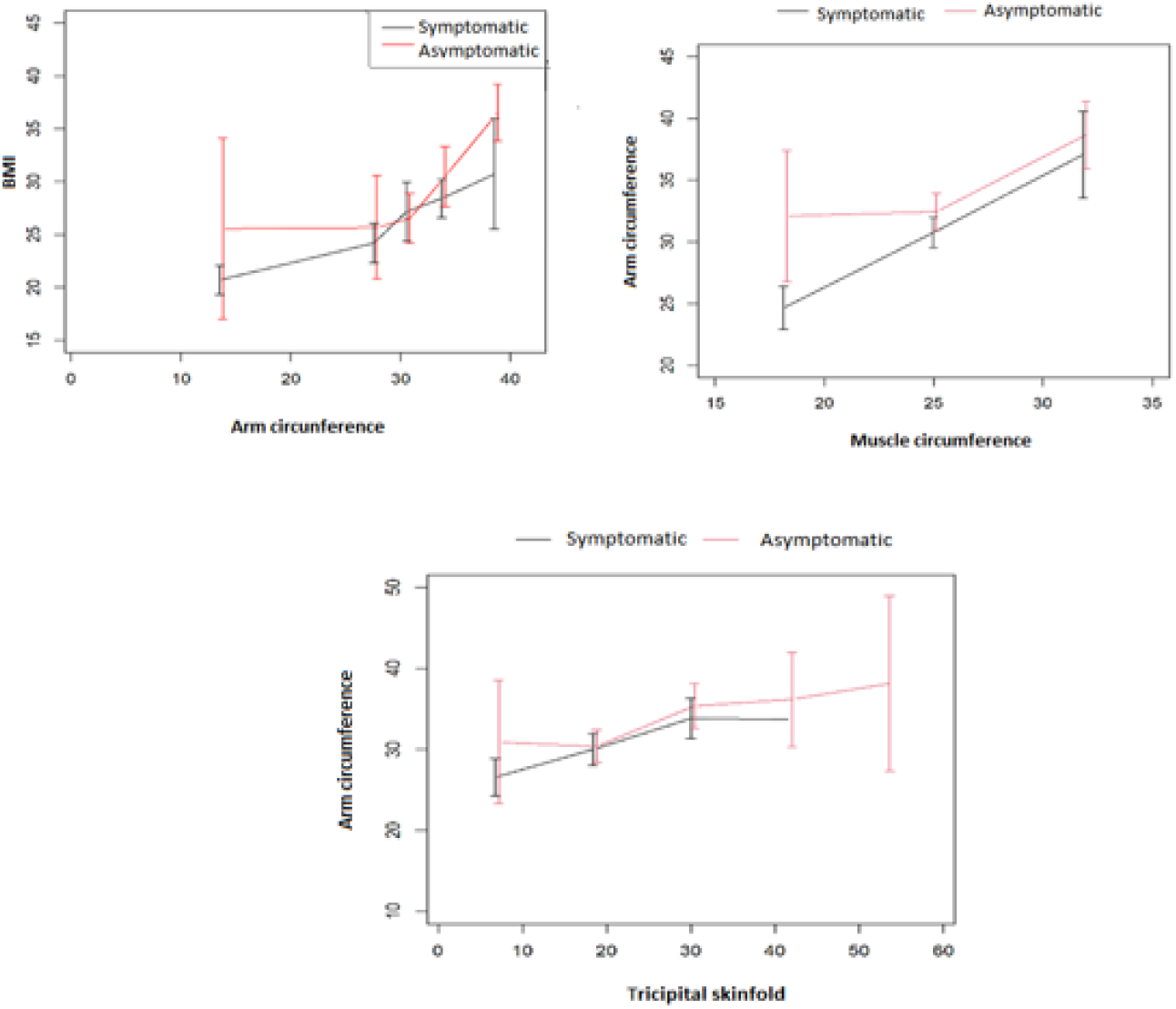
Comparison of the anthropometric parameters between symptomatic and asymptomatic HTLV. (Vertical bars are means and its 95% confidence interval of aggregate subset values).

The mean PA is slightly lower in the symptomatic group. Additionally, the same patterns can be observed at most of the strata of sex and age, with a most evident difference between groups of older females. In general, corroborating the BMI interpretation, the asymptomatic group has lower mean %LM values and higher %FM mean values. In the latter case it is more evident in the older male strata and regression (Table 3 and Figure 3).

**Table 3.** Bioelectrical Impedance Analysis (BIA) for HTLV clinical status stratified by age and sex.

| Variable | Sex<br>Female/ Male | Age<br>Years | Symptomatic<br>Mean ±SD (min,max) [N] | Asymptomatic<br>Mean ±SD (min,max) [N] | Total Mean ±SD (min,max) [N] | P. value |
| --- | --- | --- | --- | --- | --- | --- |
| Lean mass (%) | Female | 27-59 | 61.54 ±12.36 (56.55,67.70) [16] | 57.75 ±2.18 (56.85,57.83) [8] | 60.28 ±10.22 (56.85,65.12) [24] | 0.1332 |
| | Female | 60-81 | 66.49 ±8.10 (62.33,69.83) [16] | 64.79 ±11.44 (59.05,70.80) [15] | 65.67 ±9.73 (59.35,70.75) [31] | 0.5453 |
| | Male | 27-59 | 68.85 ±15.60 (68.50,76.53) [14] | 73.17 ±4.31 (70.50,76.12) [4] | 69.81 ±13.88 (68.50,76.58) [18] | 0.7980 |
| | Male | 60-81 | 75.95 ±6.14 (71.45,79.20) [12] | 68.86 ±4.10 (66.70,72.80) [5] | 73.86 ±6.42 (68.40,76.70) [17] | 0.0365 |
| Fat (%) | Female | 27-59 | 35.02 ±12.27 (28.70,38.62) [16] | 42.25 ±2.18 (42.17,43.15) [8] | 37.43 ±10.57 (33.38,42.90) [24] | 0.0131 |
| | Female | 60-81 | 34.14 ±6.57 (30.18,37.67) [16] | 35.21 ±11.44 (29.20,40.95) [15] | 34.65 ±9.11 (29.25,40.65) [31] | 0.5717 |
| | Male | 27-59 | 29.44 ±10.11 (23.48,31.50) [14] | 28.57 ±3.11 (27.62,29.95) [4] | 29.24 ±8.95 (24.45,31.50) [18] | 0.7209 |
| | Male | 60-81 | 23.43 ±6.79 (19.35,28.55) [12] | 31.14 ±4.10 (27.20,33.30) [5] | 25.70 ±7.00 (23.30,31.60) [17] | 0.0365 |
| Phase angle (PA) | Female | 27-59 | 5.77 ±1.24 (5.00,6.55) [16] | 6.53 ±0.74 (5.95,7.00) [8] | 6.02 ±1.14 (5.47,6.78) [24] | 0.1979 |
| | Female | 60-81 | 4.98 ±1.03 (4.55,5.55) [16] | 6.00 ±1.50 (5.10,6.60) [15] | 5.47 ±1.36 (4.75,6.15) [31] | 0.0326 |
| | Male | 27-59 | 5.99 ±1.01 (5.53,6.47) [14] | 6.75 ±0.44 (6.50,6.80) [4] | 6.16 ±0.96 (5.62,6.57) [18] | 0.0789 |
| | Male | 60-81 | 6.40 ±1.05 (5.83,6.62) [12] | 5.74 ±0.79 (5.20,6.00) [5] | 6.21 ±1.00 (5.40,6.60) [17] | 0.2249 |

Max= Maximum, Min=Minimum, N= sample size, SD = Standard Deviation, %= Percentage; p value for the wilcox test

**Figure 3.**
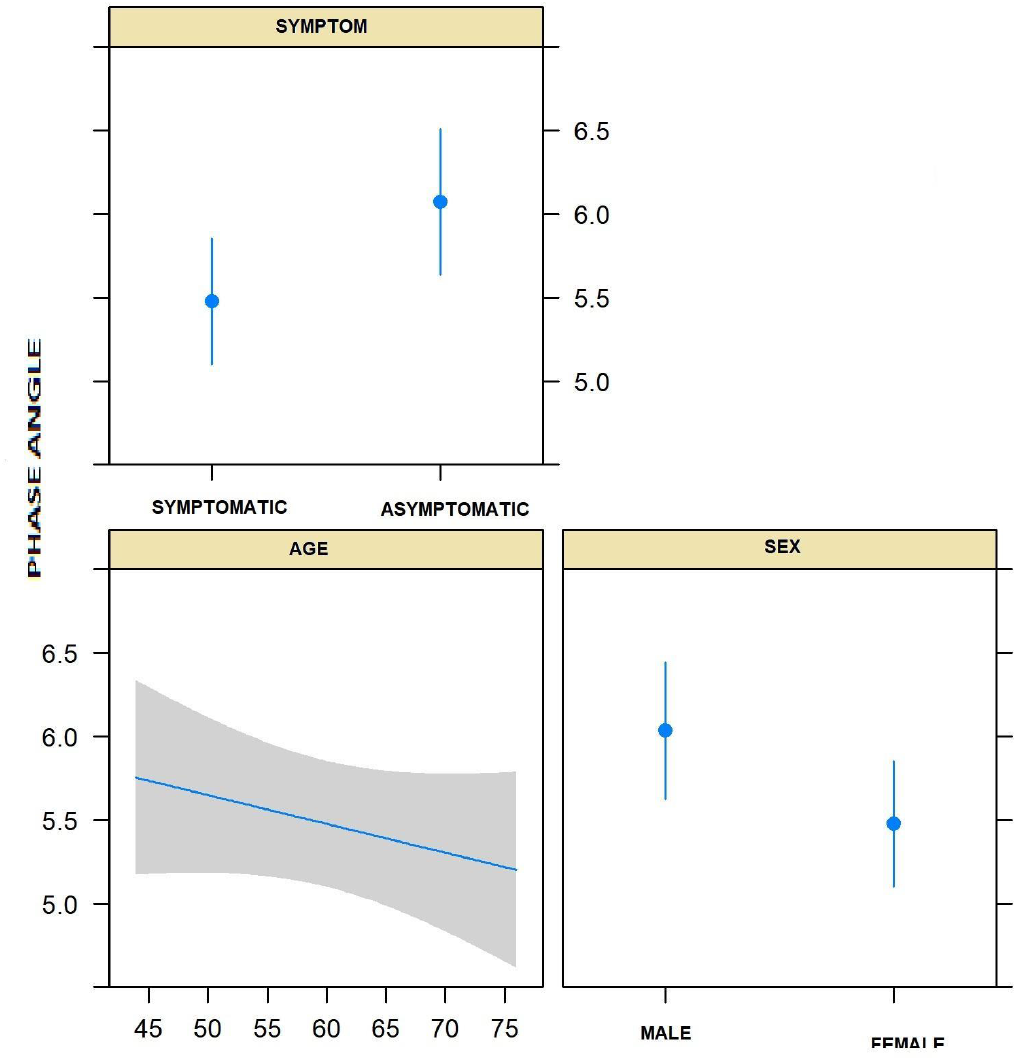
Linear regression adjusting the phase angle by symptomatology, age and sex.

When adjusting the effect of HTLV symptoms, for age and sex on PA (Figure 3), it seems the the PA deacreses slighly while age increases, female and symptomatic groups have slightly lower meas effect on PA values but none of these states reached

## DISCUSSION

The main findings of this study indicate that: a) BMI, MUAC and %FM were lower in symptomatic than in asymptomatic patients; b) symptomatic group had a lower proportion of overweight/obesity; c) MUAC can be considered an appropriate marker to replace BMI in monitoring the nutritional status of patients with HTLV-1; d) all patients which HTLV showed PA values below the cut-off point, suggesting damage to the membrane integrity in both groups.

The literature regarding nutritional status of patients with HTLV-1 is scarce. This was one of the few studies that described in detail the nutritional status of symptomatic and asymptomatic people infected with HTLV-1. This study demonstrates that overweight and obesity were prevalent in these HTLV patients, more evident among the asymptomatic group, similarly to the situation observed in the general population, corroborating what was observed in the few studies available (23–25).

As the symptomatic group has lower BMI, MUAC and %FM, and a lower frequency of overweight and obesity, it is possible that chronic HTLV infection may lead to a progressive and constant inflammatory response, increasing the energetic protein catabolism (26).

This relation gains potential attention in this study since it was possible to identify that BMI, MUAC, and the %FM were lower and the lean % LM was higher in symptomatic individuals, who need assistive devices for displacement, than in asymptomatic individuals. Therefore, although there seems to be a mobilization of lean mass, we can suggest that there is also a large mobilization of fat mass. It is noteworthy that the limitation of physical mobility to walk may have contributed to the preservation of the muscle mass of upper limbs by physical stimulus with the use of crutches and wheelchairs, which increases the uptake of amino acids by the tissue (27).

Thus, we are led to the discussion about the possibility of exchanging the proportion of fat mass for lean mass in the symptomatic population, as a way of adapting to the condition of need for strength for physical mobility (28). Considering that symptomatic may require assistance to walk, such as walking stick or wheel chair, increased values in MUAC suggests that the impact of some possible physical limitations in the lower limbs creates the need for greater effort in the upper limbs; while an asymptomatic population tends to be more sedentary (27,29).

The use of BMI may be harmed in populations with limited walking and orthostatic positioning. In this setting, adequate equipment for reliable verification may be required. However, it is satisfactory to identify that in the studied population, there was an agreement between the measures of MUAC and BMI.

MUAC was previously suggested in elderly or in hospitalized patients to monitor nutritional status (30,31). This evidence came up through the difficulties in verifying weight and height in a low mobility population. An anthropometric tape is enough for MUAC’s verification, which seems to be an advantage over BMI due to price and easier assessment (29,32,5). It is reasonable to suggest MUAC as a protocol for monitoring nutritional status in HTLV-1, as it is an anthropometric indicator that can be used longitudinally and it is correlated to other nutritional status biomarkers (32).

This study does not aim to describe the occurrence of metabolic changes associated with weight gain in the HTLV-1 population. Nonetheless, chronic metabolic diseases are positively related to being overweight, and this condition contributes to increased inflammation (33,34).

The chronic inflammation impact on infectious diseases population is still unknown; mainly considering the metabolic impact of stress induced by the viral infection itself, which associated with senescence, as cellular aging, can trigger organic failure related to degenerative diseases (35).

Therefore, the finding of higher BMI and MUAC in asymptomatic individuals, and its relationship with the highest %FM, suggests the use of adipose reserves and the preservation of LM, similar to that found in other studies with and without the disease (32,35).

The %LM was proven to be a good indicator of nutritional status for this population, corroborating studies that state that the BIA should be done whenever possible in order to guarantee the correct interpretation of the individual’s nutritional status (32). As well as the finding of a %LM inversely proportional to BMI and TSF, as well as the idea that the increase in MUAC and BMI in HTLV-1 patients tend to be the result of adipose tissue is corroborated by studies of Bacelo (22).

The finding of a lower percentage of lean mass in symptomatic older men suggests that senescence, a condition that promotes less able to preserve muscle tissue, is potentiated in the male population (36).

As seen in this study, Barbosa Silva (2005) found no significant statistical difference regarding PA (33); but, it is important to describe that in our studies the relationship between PA and variables such as gender, age, race, and other body composition indicators suggest a difference in symptomatic males and in adult females. Which is extremely important, mainly, considering that higher PA values are related to better cell membrane integrity (37).

Another important finding was that, regardless of symptoms, we found a predominance of overweight and obesity associated with PA. Bosy-Westphal (2006) (38), who obtained a score between BMI and PA in qualifying in their results the young population with lower BMI values in people living with obesity, but in our study the younger are living with obesity with greater PA, and the older male population with symptoms showed the best BMI.

It suggests that although it could be still early for any prognostic proposal for a capacity of PA in this population, the systemic condition of the chronic infection points out that: a) HTLV-1 infection alone is already capable of generating damage to the cell membrane; b) the clinical evolution of the disease when associated with senescence can aggravate the damage in the integrity of the cellular membrane, mainly in women, c)patients, even asymptomatic, should be considered at nutritional risk.

## LIMITATIONS

Our main limitation was the impossibility of a biochemical analysis in this group, that could clarify the relationship between nutritional status and an eventual metabolic illness. But it was not the object of our study.

The aim of the study was to compare the FA in symptomatic and asymptomatic patients with HTLV, we aim in future studies to compare this population with a control group. Since, there is an intrinsic limitation in its use regarding its interpretation in different settings or populations (14).

## STRONG POINTS

The main strong point of this study was comparing body composition and population discrimination between symptoms and asymptomatics and larger sample size than other nutrition studies with HTLV-1 participants.

## CONCLUSIONS

Based on the data, it can be concluded that overweight and obesity were predominant. However, the asymptomatic group is more obese than the symptomatic group. The arm’s circumference proved to be a good parameter to be used in the population with HTLV-1. The phase angle cannot be used as a prognosis because it is already altered in asymptomatic patients. This finding suggests that HTLV-1 infection alone can be considered a nutritional risk and may promote damage to the membrane integrity of asymptomatic HTLV-1 patients. So, further studies on the correlation between the infectious conditions are needed.

## Data Availability

If the data are all contained within the manuscript and/or Supporting Information files, enter the following: All relevant data are within the manuscript and its Supporting Information files.
This study was approved by the Research Ethics Committee of the National Institute of Clinical Infectious Diseases Evandro Chagas under number CAAE 46029415.9.0000.5262 on 09/14/2015

## AKNOWLEDGEMENTS

We thank the support of the nutrition service, National Institute of Infectious Diseases Evandro Chagas - Oswaldo Cruz Foundation the Research Support Foundation of the State of Rio de Janeiro (Faperj) for the financing and the neuroinfection laboratory - INI Fiocruz for the development of this study. Finally, our most sincere thanks to all the volunteers who agreed to participate voluntarily in making this study feasible.

## DECLARATIONS OF AUTHORSHIP

The data collection was extracted and checked by Rocha and Torrez, reviewed by Bacelo, Almeida and Brito. Rocha wrote the first version of the article, which Araújo and Fonseca initially reviewed. Cople-Rodrigues and Fonseca highlighted the most relevant articles. Rocha, Brito and Bacelo did the second review. Rocha, Fonseca and Cople-Rodrigues did the third review. Marcel contributed to the beginning of the statistical analysis, Brasil revised and completed the statistical analysis. Bacelo, Araújo and Cople-Rodrigues reviewed the latest versions of writing construction until the final version. All authors contributed to the construction of the article. All authors declare to have no conflict of interest.

## ABBREVIATIONS

AC: Arm Circumference
AMC: Arm Muscle Circumference
ATL: Leucemia/linfoma de células T
BIA: Bioimpedance
BMI: Body Mass Index
ELISA: Enzyme Linked Immuno Sorbent Assay
FM: Fat mass
HTLV: Human T-cell Leukemia-lymphoma Virus
INI Fiocruz: (Instituto Nacional de Infectologia, in portuguese) Evandro Chagas National Institute of Infectious
LBM: Lean Body Mass
LM: Lean mass
MUAC: Mid-arm muscle circumference
OMS: World Health Organization
PA: Phase Angle
PEM: Protein-energy malnutrition
R: Resistance
SISVAN: Vigilância alimentar e nutricional (Food and Nutrition Surveillance System)
TCLE: Termo de Consentimento Livre e Esclarecido (Free and Informed Consent Term)
TSF: Tricipital Skinfold
TSP: Tropical spastic paraparesis
TSP/HAM: Tropical spastic paraparesis / Myelopathy
XC: Reactance

